# SARS-CoV-2 seroprevalence among public school staff in Metro Vancouver after the first Omicron wave in British Columbia, Canada

**DOI:** 10.1101/2022.07.04.22277230

**Authors:** Allison W Watts, Louise C Mâsse, David M Goldfarb, Michael A Irvine, Sarah M Hutchison, Lauren Muttucomaroe, Bethany Poon, Vilte E Barakauskas, Collette O’Reilly, Else S Bosman, Frederic Reicherz, Daniel Coombs, Mark Pitblado, Sheila F O’Brien, Pascal M Lavoie

## Abstract

**Objective:** To determine the SARS-CoV-2 seroprevalence among school workers in the setting of full in-person schooling and the highly transmissible Omicron variants of concern.

**Design:** Cross-sectional study among school staff, comparing to period-, age-, sex- and postal code-weighted data from Canadian blood donors from the same community.

**Setting:** Three large school districts in the greater Vancouver metropolitan area, British Columbia, Canada, with serology sampling done between January 26, 2022 and April 8, 2022.

**Participants:** School staff actively working in the Vancouver, Richmond and Delta School Districts.

**Main outcome measure:** SARS-CoV-2 seroprevalence based on nucleocapsid (N)-protein testing, adjusted for the sensitivity and specificity of the assay.

**Results:** A majority (65.8%) of the 1845 school staff enrolled reported close contact with a COVID-19 case outside the household. Of those, about half reported close contact with a COVID-19 case at school either in a student (51.5%) or co-worker (54.9%). In a representative sample of 1620 (87.8%) school staff, the adjusted seroprevalence was 26.5% [95%CrI: 23.9 – 29.3%]. This compared to an age, sex and residency area-weighted seroprevalence of 32.4% [95%CrI: 30.6 – 34.5%] among 7164 blood donors.

**Conclusion:** Despite frequent COVID-19 exposures, the prevalence of SARS-CoV-2 infections among the staff of three main school districts in the Vancouver metropolitan area was no greater than a reference group of blood donors, even after the emergence of the more transmissible Omicron variant.

**What is already known on this subject?:**

- Earlier studies indicate that COVID-19 infection rates are not increased among school staff at previous stages of the pandemic compared to the community, yet controversy remains whether this will remain true after the emergence of the highly transmissible Omicron variant.

**What this study adds?:**

- Despite frequent COVID-19 exposures, this study identified no detectable increase in SARS-CoV-2 seroprevalence among school staff working in three metro Vancouver public school districts after the first Omicron wave in British Columbia, compared to a reference group of blood donors from the same age, sex and community area.

## Introduction

The COVID-19 pandemic has had significant impact on the school system, causing major stress and disruptions with mental and physical consequence on students and staff ^1^. The risk of secondary viral transmission within the school setting has been intensely debated. While SARS-CoV-2 outbreaks have occurred, contact tracing studies while mitigation measures were in place (e.g., masking, symptoms checks) have generally shown that the majority of student and staff cases originate from the community rather than within schools ^2-12^, and that the overall risk of infection among school staff remains very low ^13-16^. Supporting these data, SARS-CoV-2 infections within schools in Canada ^11 17 18^ as well as the UK and Europe ^13 19 20^ have also been reported to be low overall during the 2020-2021 school year. However, most of these studies were conducted before the emergence of highly transmissible variants.

At the end of 2021, a new SARS-CoV-2 BA.1 (Omicron) variant, that was first reported in South Africa, began to emerge across the world ^21 22^, rapidly replacing other strains and causing a massive wave due to its highly transmissible nature and ability to evade vaccine protection against re-infection ^23 24^.

In British Columbia (Canada), relatively low rates of COVID-19 had been reported in the earlier phases of the pandemic compared to other areas. By the end of the spring 2020, a very low proportion (∼1%) of people living in Vancouver, BC had been infected with SARS-CoV-2 ^25-27^. By March 2021, about 2-5% of school staff among the Vancouver public school system showed evidence of a prior SARS-CoV-2 infection, based on antibody testing ^17^. At the end of 2021, the world experienced a massive increase in cases, with the emergence of the highly transmissible Omicron variant ^28 29^. This rise in COVID-19 cases dramatically heightened levels of stress expressed in the public space, particularly among school workers ^30 31^. As new cases also exceeded the testing capacity of health systems, BC re-prioritized viral testing towards selected groups around January 17, 2022. Given this change in wide access to testing, it became nearly impossible to publicly and accurately track the total number of cases in the community during and subsequent to the Omicron wave, and thus, the transmission of SARS-CoV-2 within schools.

The main objective of this study was to determine the seroprevalence of SARS-CoV-2 infections among the school staff of three of the largest school districts within the greater Vancouver area, after the first Omicron wave in British Columbia and Canada. The second objective was to compare infection rates among school workers to the community rate, based on blood donor data collected over the same period, age, sex and residential area.

## Materials and Methods

### Study design and participants

This study is a cross-sectional analysis of data collected from staff working in three school districts (Vancouver, Richmond and Delta) within the greater Vancouver Area. The study included current, full or part-time district staff members recruited from a prospective longitudinal study conducted from February to June 2021^17^. Informed consent was obtained. The study excluded staff who were: temporary, on-leave, or on-call with no classroom time, or working exclusively in adult education.

For this study, staff who participated in the 2021 seroprevalence study (N=2538) ^17^ were emailed again on January 26, 2022, with subsequent reminder emails in the following weeks. Recruitment ended on March 31, 2022. Blood samples were collected for serology between January 27 and April 8, 2022, shortly after the first Omicron wave in BC, (**Supplemental Figure 1**). Comparative community data were obtained from Canadian blood donors who did not have COVID-19 at the time of serology, between January 1 and March 31, 2022. All study procedures were approved by the University of British Columbia Children’s and Women’s Research Ethics Board (H20-03593).

### Study setting

The Vancouver, Richmond and Delta school districts include 151 elementary and 18 secondary schools distributed in the greater metropolitan area of Vancouver, BC, Canada. Together, they serve a population of ∼935,000 (2.6 million people live in the greater urban area). During the 2021-2022 school year, schools remained open as usual. COVID-19 mitigations measures implemented in district schools and indications for viral testing are detailed in **Appendix 1**.

### Data collection

Data on demographic factors, occupation, health status and history of COVID-19 exposure (e.g., testing, vaccination, masking behaviors) were collected from the school staff via an online questionnaire ^32^. An additional questionnaire which asked about mental health is not reported in this paper. Blood donors were asked their age, sex and postal code of residence at the time of blood donation using questionnaires administered by Canadian Blood Services as part of the routine donation process.

### Serology testing

Blood samples were collected at clinics set-up in participating Vancouver schools, at the BC Children’s Hospital or outpatient clinical laboratories in the greater Vancouver area. Testing for anti-nucleocapsid (N) protein SARS-CoV-2 antibodies was performed using the Health Canada and FDA-licensed qualitative total antibody Roche Elecsys^*TM*^ Anti-SARS-CoV-2 anti-nucleocapsid assay (Roche, USA). Testing was performed on a Cobas e601 analyzer at Canadian Blood Services national lab in Ottawa, Canada. Specimens were considered reactive at a cut-off index ≥1.00. N antibodies have been shown to persist in blood with assay sensitivity maintained until at least a year post-infection ^33^.

### Bias minimization strategies

To ensure recruitment bias was minimized, we developed an active recruitment and facilitation strategy where district leaders, Teacher and Student Support Worker, and parent Associations were engaged right from the study design stage. Weekly meetings occurred from study launching until publication with a Vancouver school district leadership representative and a district liaison. For serology sampling, blood collection sites were set-up in a variety of geographically dispersed schools over lunch and after work or participants could attend one of over a hundred private community clinics open on weekends or the British Columbia Children’s Hospital (for collection both within and outside normal working hours). A full-time study coordinator was hired to maintain contact with participants 7 days per week, facilitate blood collection with flexible hours, including driving around the city to meet the few participants who were unable to attend the blood clinics. Participants received a $20 incentive and their serology results. Serology testing among school staff was evenly distributed across Vancouver district schools with high and low numbers of reported COVID-19 cases (data not shown).

### Statistical analyses

Descriptive statistics were run using STATA version 17.0. To examine differences in seroprevalence across school type and occupation, logistic regression models using the margins command were run to obtain the seroprevalence and 95% confidence intervals (95%CI) for each subgroup. A Bayesian analysis was conducted incorporating a hierarchical design to account for variation in prevalence between school districts. For this analysis, 182 out of 205 positive controls (based on our own unpublished data in a fully vaccinated adult population), and 10432 out of 10453 negative controls (based on data from the manufacturer, at: https://diagnostics.roche.com/global/en/products/params/elecsys-anti-sars-cov-2.html) were incorporated using a binomial likelihood to account for the true evidence of test sensitivity and specificity. Weakly informative priors were selected for the baseline prevalence, logit sensitivity, logit specificity, and scale of the regional variance. 2000 samples were generated from the posterior with 1000 warm-up iterations and no thinning across 4 chains using the NUTS sampling algorithm. Effective sample size, Gelman-Rubin statistic, and visual inspection of posterior sample chains were used to determine convergence, mixing, and adequate sample size. Results were reported using the expectation under the posterior with 95% credible intervals (95% CrI). Bayesian analyses were conducted in R version 4.1.0 and Stan version 2.21.1. All analyses were done on cases with complete data.

### Data statement

De-identified data will be made available through the COVID-19 Immunity Task Force.

## Results

### Characteristics of school staff sample

In total, 1845 school staff were enrolled in the study (**Figure 1**). About 80.7% were classroom workers, for a median contact time with students of 18.0 [IQR: 6.0 to 30.0] hours per week. Almost all staff (98.9%) had received at least two doses of a Health Canada-approved COVID-19 vaccine. The baseline characteristics of the enrolled school staff were found to be representative of the corresponding school district populations in terms of age, sex, and residence area distribution, except for a higher cumulative incidence of COVID-19 and higher proportion of classroom and elementary staff in the school sample compared to the district populations (**Supplemental Table 1**).

**Figure 1.**
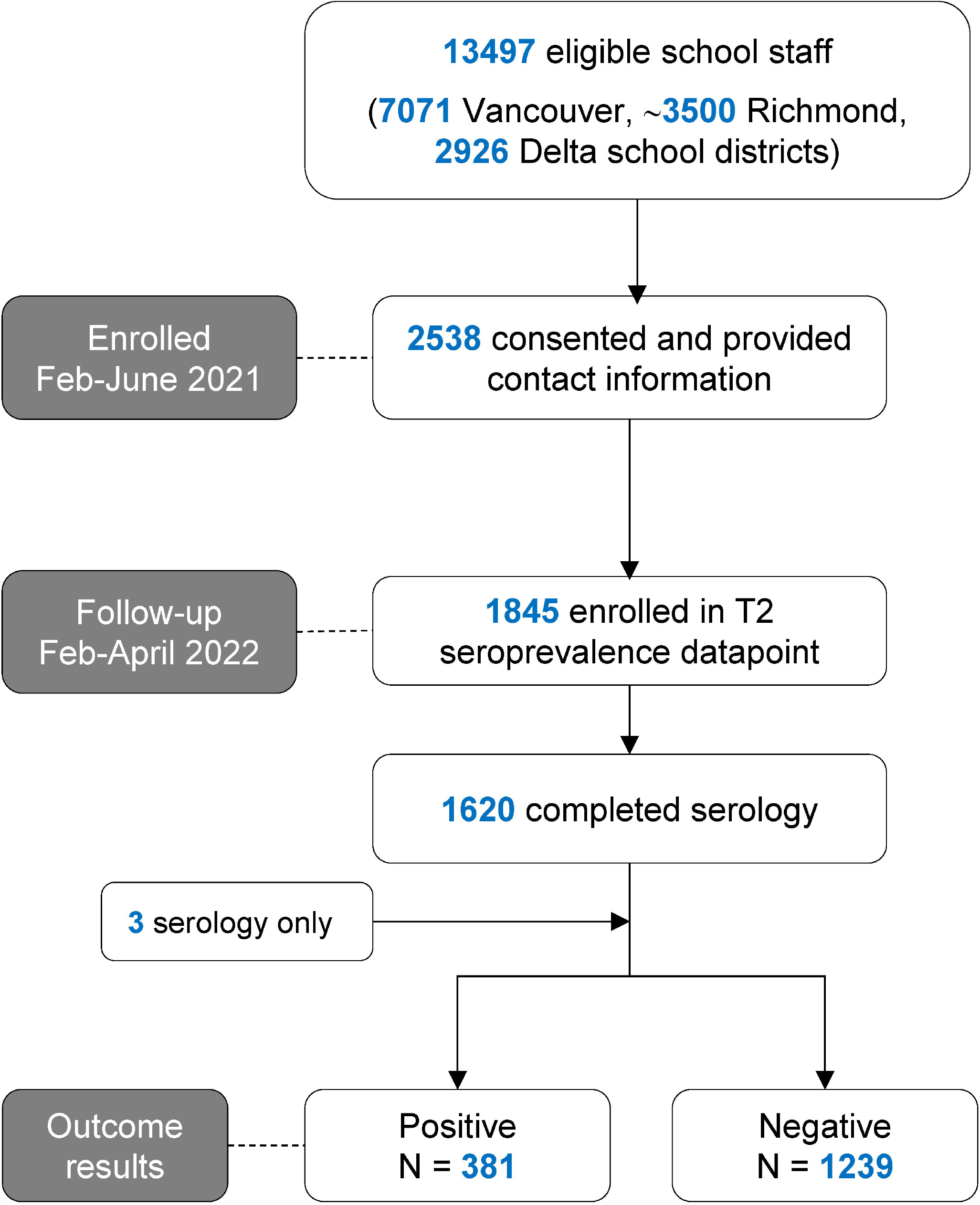
Flow diagram for enrollment of school staff study sample.

### COVID-19 exposures among school staff

The potential sources of contact with COVID-19 cases among school staff are shown in **Table 1**. About a third reported living with an essential worker, 41.1% had children and 20.7% reported ever having a COVID-19 case in the household (**Table 1**).

**Table 1:** COVID-19 exposures among school staff sample.

| **Variable** | **n<sup>#</sup>** | **School staff**<br>**(n = 1845)** |
| --- | --- | --- |
| COVID-19 case among household members<sup>δ</sup>, % Yes, n (%) | 1845 | 382 (20.7%) |
| Type of occupation for essential worker living in household, n (%) | 1845 | 662 (35.9%) |
| Agriculture & food production |  | 11 (1.7%) |
| Community services (waste disposal, emergency response, postal) |  | 45 (6.8%) |
| Construction, maintenance, skilled trades |  | 89 (13.4%) |
| Consumer products (hardware, safety, vehicle, sales, garden centres) |  | 11 (1.7%) |
| Financial services (banking, real estate, insurance) |  | 20 (3.0%) |
| Food (grocery, convenience, liquor, restaurant) |  | 68 (16.8%) |
| Health care |  | 111 (16.6%) |
| Social services, education, research |  | 287 (43.4%) |
| Manufacturing, resources, energy, utilities |  | 19 (2.9%) |
| Services (pharmacy, gas station, delivery, funeral, vet, etc.) |  | 22 (3.3%) |
| Sports (professional) |  | 5 (0.8%) |
| Supply chain & transportation |  | 37 (5.6%) |
| Telecommunications & IT (including the media) |  | 11 (1.7%) |
| Other |  | 1 (0.2%) |
| Close contact<sup>*</sup> with a COVID-19 case outside household, n (%) | 1845 | 1214 (65.8%) |
| Another school staff member / work colleague |  | 666 (54.9%) |
| Student in classroom setting |  | 625 (51.5%) |
| Friend |  | 553 (45.6%) |
| Family (non-household member) |  | 417 (34.4%) |
| Other |  | 93 (7.7%) |
| Wear a mask in public places<sup>τ</sup>, % Always or Often, n (%) | 1839 | 1824 (99.2%) |
| Co-workers wear masks<sup>τ</sup>, % Always or Usually, n (%) | 1826 | 1776 (97.3%) |
| Students wear masks<sup>τ</sup>, % Always or Usually, n (%) | 1702 | 1502 (88.3%) |
| Elementary | 1050 | 897 (85.4%) |
| Secondary | 569 | 536 (94.2%) |

<sup>#</sup>N with data available.
<sup>\*</sup>Reported contact with a known COVID-19 case within 2 meters and for >2 minutes.
<sup>δ</sup>“Since the last survey, has anyone in your household (not counting yourself) ever tested positive for COVID-19? ([Yes], [Not applicable, I live alone], [No one has been tested], [No, they tested negative], [Not sure, waiting for the result])”.
<sup>τ</sup>Questions about masking were as follows: “How often have you worn a mask in public places in the past three months? (Never, Rarely, Occasionally, Often, Always)”; “To the best of your knowledge, how often do your co-workers wear a mask in your presence? (Never, Occasionally, Usually, Always)”; “To the best of your knowledge, how often do students in your school wear a mask in your presence? (Never, Occasionally, Usually, Always)”.

Most school staff reported that co-workers and students wore masks often or always at school in the past 3 months (**Table 1**). A majority (65.8%) of the school staff reported close contact with a COVID-19 case outside the household during the pandemic, of which about half of these reported close contacts were with a COVID-19 case at school either in a student (51.5%) or co-worker (54.9%) (**Table 1**). Just over half (52.2%) of school staff reported no close contact with a school case.

About 16.3% of school staff felt they had ever had COVID-19 (**Table 2**). The cumulative incidence of COVID-19 diagnosed by nucleic acid or rapid antigen testing, self-reported among the 1845 school staff was 15.8% (**Table 2**). One school staff required hospitalization for COVID-19 (**Table 2**).

**Table 2:** Cumulative incidence of COVID-19 among school staff.

| **Variable** | **n<sup>#</sup>** | **School staff**<br>**(n = 1845)** |
| --- | --- | --- |
| Reported having COVID-19 symptoms<sup>*</sup>, n (%) | 1845 | 301 (16.3%) |
| Number tested for COVID-19 (PCR), n (%) |  | 766 (41.5%) |
| At least one positive COVID-19 PCR test |  | 120 (6.5%) |
| More than one positive COVID-19 PCR test |  | 4 (0.2%) |
| All negative COVID-19 PCR tests |  | 639 (35.0%) |
| Did not know/could not remember test result |  | 33 (1.8%) |
| Number tested +ve by Rapid Test, n (%) | 1842 | 206 (11.8%) |
| Number tested +ve by any test, n (%) | 1845 | 291 (15.8%) |
| Hospitalized for COVID-19, n (%) |  | 1 (0.01%) |

<sup>#</sup>N with data available.
<sup>\*</sup>Participants who responded that they had COVID-19 symptoms, in response to: “Do you think you’ve had covid? (Yes/No) If yes, why?”

### SARS-CoV-2 seroprevalence

In total, 1620 (87.8%) school staff underwent serology testing with a median date of testing of February 14, 2022. The characteristics of the 1620 school staff that completed serology testing were representative of all 1845 school staff in the study (**Table 3**).

**Table 3:** Characteristics of school staff sample.

| **Variable** | **n<sup>#</sup>** | **Completed questionnaire**<br>**(n = 1845)** | **n<sup>#</sup>** | **Completed serology testing**<br>**(n = 1620)** |
| --- | --- | --- | --- | --- |
| Age (mean ± SD) | 1845 | 46.7 ± 10.2 | 1620 | 47.5 ± 10.1 |
| Sex, % female, n (%) | 1837 | 1508 (82.1%) | 1616 | 1331 (82.4%) |
| Canadians of indigenous origin, n (%) | 1840 | 33 (1.8%) | 1617 | 27 (1.7%) |
| Ethnicity, n (%) | 1845 |  | 1620 |  |
| White, Caucasian |  | 1299 (70.4%) |  | 1118 (69.2%) |
| South Asian |  | 68 (3.7%) |  | 59 (3.7%) |
| Chinese |  | 309 (16.8%) |  | 288 (17.8%) |
| Black |  | 10 (0.5%) |  | 8 (0.5%) |
| Filipino |  | 31 (1.7%) |  | 28 (1.7%) |
| Latin American |  | 27 (1.5%) |  | 22 (1.4%) |
| Arab |  | 5 (0.3%) |  | 3 (0.2%) |
| Southeast Asian |  | 31 (1.7%) |  | 28 (1.7%) |
| West Asian |  | 1 (0.2%) |  | 3 (0.2%) |
| Korean |  | 14 (0.8%) |  | 13 (0.8%) |
| Japanese |  | 47 (2.6%) |  | 42 (2.6%) |
| Other/no answer |  | 55 (3.0%) |  | 45 (2.8%) |
| Classroom workers*, n (%) | 1841 | 1485 (80.7%) | 1611 | 1292 (80.2%) |
| Contact time with students (hrs/wk), median (IQR) | 1839 | 18.0 [6.0-30.0] | 1609 | 17.9 [6.0-30.0] |
| Vaccination status, n (%) | 1830 |  | 1613 |  |
| Not vaccinated |  | 15 (0.8%) |  | 13 (0.8%) |
| One dose |  | 6 (0.3%) |  | 6 (0.4%) |
| Two doses |  | 249 (13.6%) |  | 213 (13.2%) |
| Three or more doses |  | 1560 (85.3%) |  | 1382 (85.6%) |
| School level, n (%) | 1845 |  | 1620 |  |
| Elementary |  | 1083 (58.7%) |  | 954 (58.9%) |
| Secondary |  | 588 (31.9%) |  | 510 (31.5%) |
| Work at multiple levels |  | 59 (3.2%) |  | 51 (3.2%) |
| School district office/other |  | 115 (6.2%) |  | 105 (6.5%) |
| Live with children <18 years, n (%) | 1845 | 758 (41.1%) | 1620 | 666 (41.1%) |
| Live with essential worker, n (%) | 1845 | 662 (35.9%) | 1610 | 572 (35.5%) |
| At least one co-morbidity<sup>†</sup>, n (%) | 1845 | 491 (26.6%) | 1620 | 434 (26.8%) |
| Smoker, n (%) | 1836 | 26 (1.4%) | 1608 | 22 (1.4%) |

<sup>#</sup> N with data available.
\* Those who reported being a Teacher, Teacher Librarian, Resource Teacher, Student Support Worker, or Family and Youth Worker in response to the question: “What is your job title? (Teacher, Teacher Librarian, Resource Teacher, Student Support Worker, Family and Youth Worker, Administrator [Principal, Vice Principal], Administrative Assistant, Maintenance Staff, School Board Office Staff, Other). And those who responded “Other” and listed student facing positions such as student aid, counsellor, speech language pathologist, etc.
¶ any the following: hypertension, diabetes, asthma, chronic lung disease, chronic heart disease, chronic kidney disease, liver disease, cancer, chronic blood disorder, immunosuppressed, chronic neurological disorder.

Of the 1620 school staff who underwent serology testing, 381 (23.5%) were positive for SARS-CoV-2 nucleocapsid antibodies. Of those who tested positive, 272 (71.4%) believed they had a prior COVID-19 infection and 194 (50.9%) reported a previous positive viral test. The unadjusted seroprevalence was 23.7% [95% CrI: 21.7 - 26.0%] among all school district staff. The unadjusted seroprevalence among all staff was also similar to the unadjusted seroprevalence in staff working in a classroom setting, and between staff working in elementary and secondary schools (**Table 4**).

**Table 4:** COVID-19 seroprevalence by school education level and occupation^1^.

|  | Frequency | Percent cases [95%CI] |
| --- | --- | --- |
| **School Level** |  |  |
| Elementary | 222 | 23.3 [20.7-26.1] |
| Secondary | 123 | 24.1 [21.0-28.4] |
| Multiple / mixed levels | 11 | 21.6 [7.8-27.9] |
| School board office | 25 | 23.8 [13.2-30.5] |
| **Occupation** |  |  |
| Classroom Worker | 306 | 23.7 [21.4-26.0] |
| Other | 73 | 22.9 [18.3-27.5] |
| **Total** | **381** | **23.5** |

<sup>1</sup>Logistic regression models examining group differences by school level and occupation were non-significant (p>0.05)

The adjusted seroprevalence was 26.5% [95%CrI: 23.9 to 29.3%] among the staff of all school districts. In comparison, the adjusted period-, sex-, age- and residency location-weighted seroprevalence was 32.4% [95%CrI: 30.6 – 34.5%] among 7164 blood donors (**Supplemental Table 2**). There were no significant differences in seroprevalence between all three school districts (**Supplemental Table 3**).

## Discussion

This study found, first, that one quarter of school workers showed evidence of a past SARS-CoV-2 infection among three of the largest school districts in the greater Vancouver area, serving approximately 83,000 students, and altogether, representing a large proportion of school staff in BC. Second, this study found that the seroprevalence among staff was not higher compared to the community, represented by a demographically similar group of blood donors. To the best of our knowledge, this study is the first to report seroprevalence estimates among school staff after the emergence of the highly transmissible Omicron variants. Findings are in keeping with data from Canada ^17 18^, and other areas of the world ^20^ before the emergence of more transmissible variants. A major strength of this study is that it utilized sensitive antibody testing that detects asymptomatic SARS-CoV-2 infections that may not have come to clinical attention, using an N-based serology assay that identifies cases for up to a year ^19^.

About one quarter of all COVID-19 cases reported in BC during this period occurred in the regional health authority where the participating school districts are located. Despite a ten-fold increase in seroprevalence compared to the previous school year (2021-2022) ^17^, these findings support that the risk of SARS-CoV-2 infection among school staff remains low, even in the context of full in-person schooling, Omicron, and when mitigation measures were applied, with high vaccine uptake. These findings contrast with the abundant COVID-19 cases reported within schools throughout Canada, Europe and the USA, and expressed in the media throughout the pandemic, that fuelled a high level of stress among the staff and students attending those schools and their families. A potential explanation for these apparent discrepancies is to consider that most school cases are acquired from social interactions occurring outside the school setting, rather than within the school setting as supported by other Canadian data ^11 34 35^.

The finding that school staff had lower seroprevalence compared to the reference group suggests that the former may be at lower risk. This may be explained by factors including potential differences in vaccination rates or timing between groups. Moreover, the credible intervals used to determine statistical significance on the seroprevalence depends on the sensitivity of the serology test used for which there is relatively little data in the context of a vaccinated population after infection with Omicron. At the very least, the lower seroprevalence observed among school staff provides strong reassurance that school staff are at similar risk of contracting COVID-19 in schools compared to the risk in the community.

This study has limitations. First, the non-random participation implies a potential volunteer bias. Supporting that this bias was negligible, the demographics of the staff sample was representative of the study population, and this is not surprising given the substantial means that were deployed to facilitate easy participation and blood sampling among school staff right where and when they work in schools. Second, blood donors may not be a reliable estimate of community seroprevalence though based on previous data they are likely more representative of school staff compared to other socioeconomic groups at higher risk of COVID-19 ^36 37^. Based on this assumption, a lower SARS-CoV-2 seroprevalence among blood donors could only reinforce our conclusions. Third, this study was conducted before mask mandates were lifted. Ongoing monitoring is warranted to determine if these conclusions will hold true as schools continue to relax mitigation measures and with the emergence of new SARS-CoV-2 variants.

In conclusion, this study confirmed that a substantial proportion (26.5%) of school staff working in three metro Vancouver public school districts showed evidence of SARS-CoV-2 infections after a major, and first Omicron wave in BC. However, the study identified no detectable increase in seroprevalence compared to a reference group of blood donors from the same age, sex and residency area. The potential waning of immunity among school staff and loosening of mitigation measures, including the lifting of mask mandates, requires ongoing evaluation of COVID-19 infections within the school community.

## Data Availability

De-identified data will be made available through the COVID-19 Immunity Task Force.

## Authors’ contributions

LCM and PML obtained funding for this study; AW, DMG, SH, MAI, DC, PML and LCM designed the study; FR reviewed the literature; LM set-up and coordinated the recruitment of participants; ESB constructed and managed the data collection database; BP processed blood samples, under the supervision of PML and VEB; MAI performed statistical analyses; SO provided matched data from Canadian blood donors; AW and MP performed data analyses; CO facilitated communications within the Vancouver District; AW & PML wrote the first draft of the manuscript. All other authors revised the manuscript and approved its final version.

## Acknowledgements

We would like to thank the school staff who participated in the study and have been working tirelessly during this pandemic, and the District leadership, particularly Suzanne Hoffman and David Nelson for providing full support during this study; Kathy O’Sullivan for ongoing work reviewing District communications, documents and liaising with school partners throughout the study; Esther Alonso-Prieto for help with the human resource and financial management of this study; Brandon Bates, John Bhullar and Chemistry Laboratory staff at Children’s and Women’s Hospitals and the BC Children’s Hospital Biobank staff for help with sample collection and processing; the District and BC Children’s Hospital communication teams, VCH’s Office of the Chief Medical Health Officer, the BC Centre for Disease Control; Steven Drews and Qi-Long Yi for consultation with data from Canadian Blood Services. We thank LifeLabs for partnering with us commercially to collect a portion of the blood samples.

