## Supplemental Files for "SARS-CoV-2 seroprevalence among public school staff in Metro Vancouver after the first Omicron wave in British Columbia, Canada"

**Supplemental Figure 1: Serology sampling in school staff in relation to COVID-19-related hospitalizations and viral Nucleic Acid Amplification Testing (NAAT) positivity rates in British Columbia - Dec 1, 2021 and June 1, 2022.**

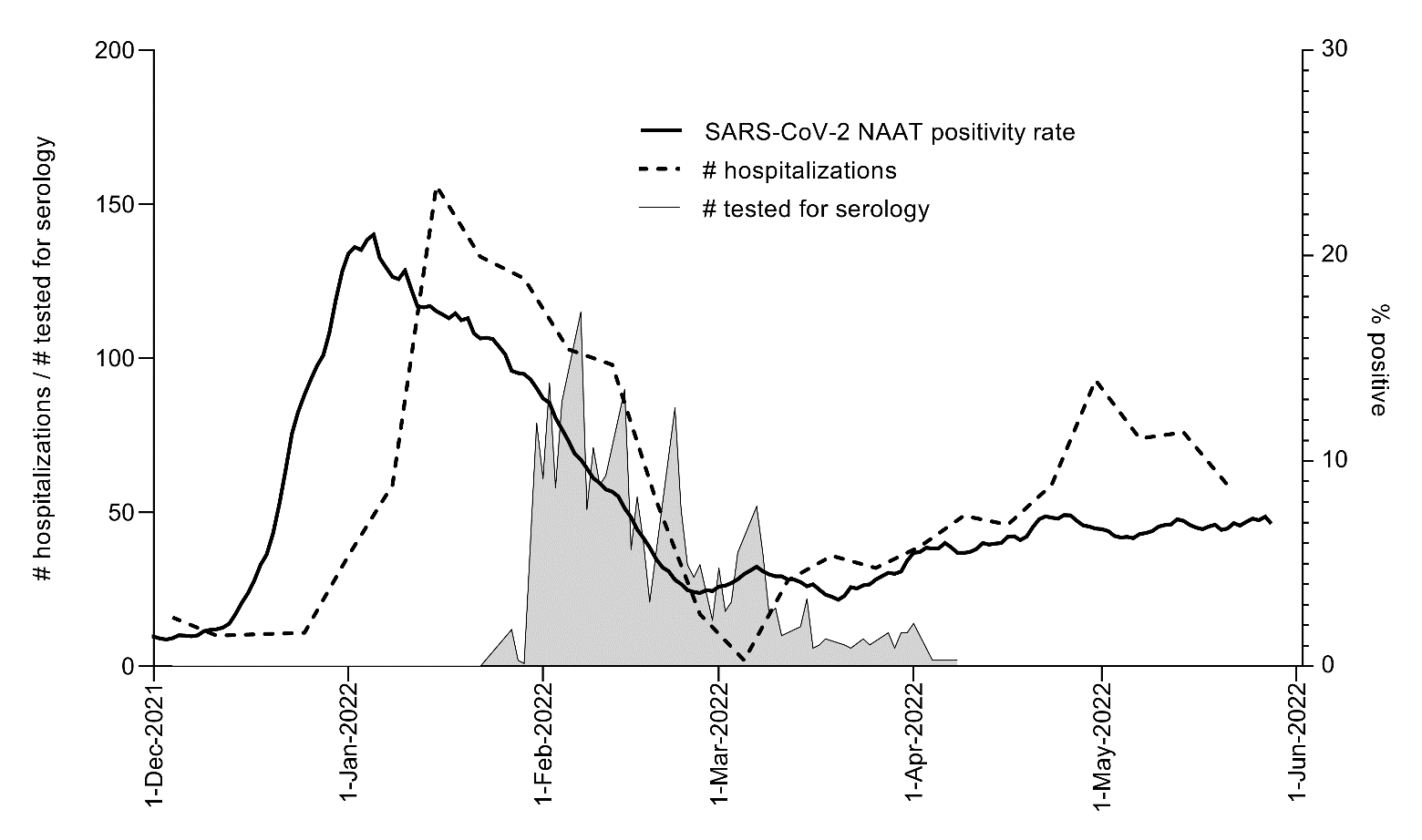

**Supplemental Figure 2: Median antibody reactivity indices among a) all school staff, b) school staff who reported a positive SARS-CoV-2 Rapid Antigen Test (RAT) and c) school staff who reported a positive Nucleic Acid Amplification Test (NAAT).** Boxes (median, with 25^th^ and 75^th^ centiles) and whiskers (min to max values). Dotted line = positivity threshold ≥ 1.00 reactivity index.

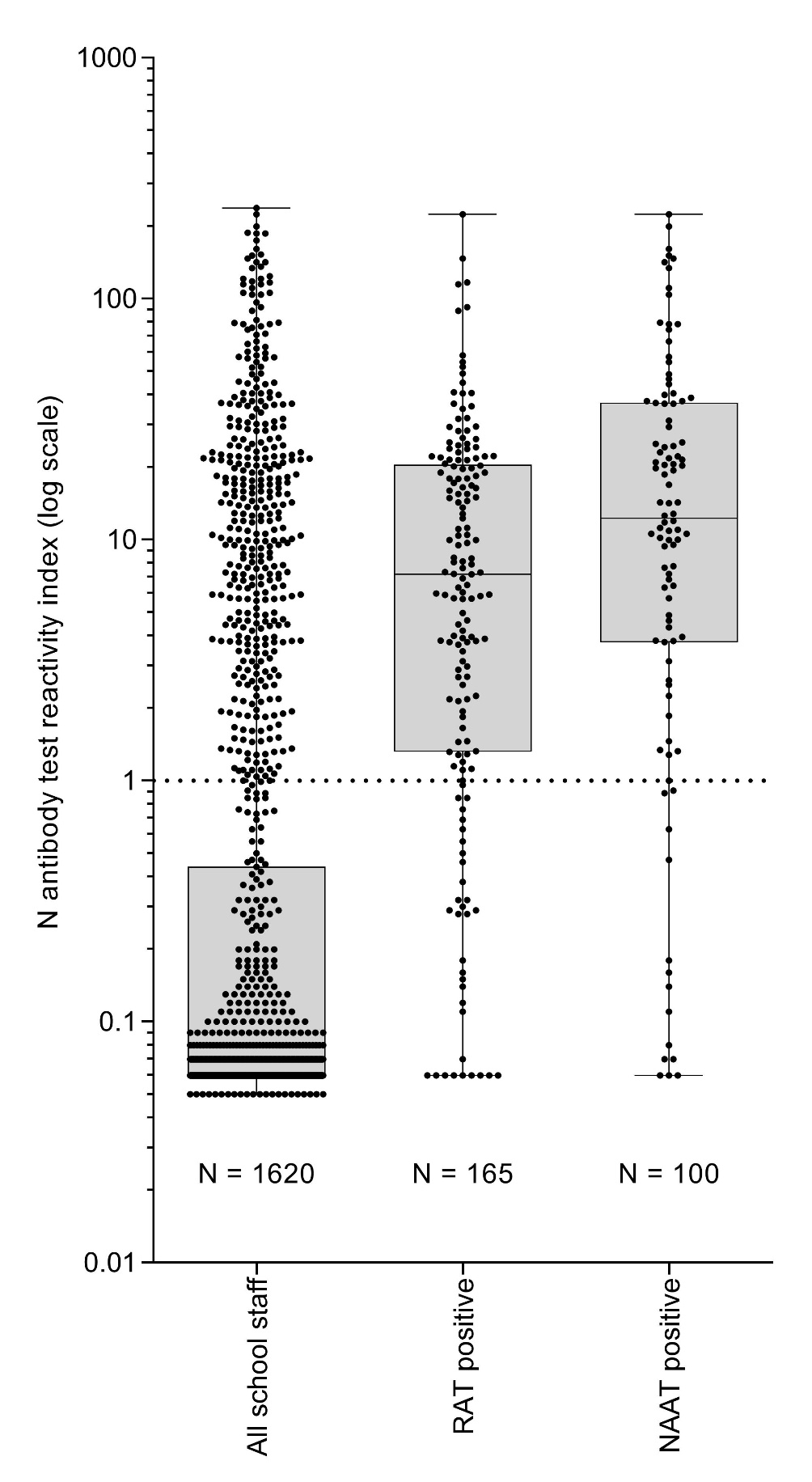

**Supplemental Table 1: Characteristics of school staff sample versus the entire corresponding school Districts**

|  | **School district** | | | | | |
| --- | --- | --- | --- | --- | --- | --- |
|  | **Vancouver** | | **Richmond** | | **Delta** | |
| **Variable** | **Staff sample**  **(n = 1277)** | **Population**  **(n = 6872)**^&^ | **Staff sample**  **(n = 305)** | **Population**  **(n ~ 3500)** | **Staff sample**  **(n = 253)** | **Population**  **(n = 2926)** |
| Age (mean ± SD) | 46.9 ± 10.1 | 47.6 ± 11.9 | 46.5 ± 10.6 | NA | 48.0 ± 10.7 | 45.3 ± 12.1 |
| Female, % | 81.1 | 68.4 | 85.2 | NA | 83.2 | 77.6 |
| School type*, % |  |  |  | NA |  |  |
| Elementary School | 65.6 | 64.7 | 61.3 |  | 64.5 | 58.9 |
| Secondary School | 34.4 | 35.3 | 38.7 |  | 35.5 | 41.1 |
| Area of residency (by 2 digits postal code), % |  |  |  | NA |  |  |
| V0 | 0.2 | 0.3 | 0 |  | 0 | 0.1 |
| V1 | 0 | 0.1 | 0.7 |  | 0 | 0.7 |
| V2 | 0.2 | 0.8 | 0 |  | 0.4 | 1.3 |
| V3 | 7.1 | 11.0 | 5.6 |  | 24.0 | 26.3 |
| V4 | 2.0 | 3.2 | 7.9 |  | 62.0 | 56.6 |
| V5 | 51.6 | 50.5 | 10.5 |  | 6.8 | 6.5 |
| V6 | 30.5 | 26.4 | 29.3 |  | 4.8 | 4.9 |
| V7 | 8.3 | 7.7 | 46.1 |  | 2.0 | 3.1 |
| V8 | 0 | 0.2 | 0 |  | 0 | 0.1 |
| V9 | 0.1 | 0.3 | 0 |  | 0 | 0.1 |
| Other | 0.1 | 0.09 | 0 |  | 0 | 0.3 |

*****of those who report working exclusively in either an elementary or secondary school; NA = Data were not be provided by school authorities for the two smaller districts; 10 participants moved to other school districts and therefore have been excluded from this table.

**Supplemental Table 2: Weighted seroprevalence among Canadian blood donors**

|  | | School staff | Blood donor community | |
| --- | --- | --- | --- | --- |
|  |  | N (%) | Unweighted N (%) | Weighted N^#^ (%) |
| Month | Jan | 94 (5.9) | 1911 (26.7) | 419 (5.9) |
|  | Feb | 1131 (70.4) | 2443 (34.1) | 5042 (70.4) |
|  | Mar | 363 (22.6) | 2520 (35.2) | 1618 (22.6) |
|  | April | 19 (1.2) | 290 (4.1) | 85 (1.2) |
| Sex | F | 1327 (82.6) | 3579 (50.0) | 4609 (82.5) |
|  | M | 280 (17.4) | 3585 (50.0) | 1248 (17.5) |
| Age (years) | 17-24 | 4 (0.25) | 61 (0.85) | 18 (0.25) |
|  | 25-39 | 367 (22.8) | 2715 (37.9) | 1636 (22.8) |
|  | 40-59 | 1059 (65.9) | 3088 (43.1) | 4721 (65.9) |
|  | ≥60 | 177 (11.0) | 1300 (18.2) | 789 (11.0) |
| FSA2 | V0 | 4 (0.25) | 92 (1.3) | 18 (0.25) |
|  | V1 | 2 (0.13) | 97 (1.4) | 9 (0.12) |
|  | V2 | 5 (0.31) | 231 (3.2) | 22 (0.31) |
|  | V3 | 140 (8.9) | 2026 (28.3) | 624 (8.7) |
|  | V4 | 176 (11.0) | 1183 (16.5) | 785 (11.0) |
|  | V5 | 634 (39.5) | 1543 (21.5) | 2826 (39.5) |
|  | V6 | 414 (25.8) | 1293 (18.1) | 1846 (25.8) |
|  | V7 | 228 (13.4) | 511 (7.1) | 1016 (14.2) |
|  | V8 | 3 (0.19) | 118 (1.7) | 13 (0.19) |
|  | Other | 1 (0.06) | 70 (0.98) | 4 (0.06) |
| Total |  | 1607* (99.2^&^) | 7164 |  |
| ROCHE N, positive | | | 1987 (27.7) | 2077 (29.0) |

^*^with data available (some staff had missing age, sex, or a residency location outside the “V” postal code).

^&^Percentage of entire school staff sample (n = 1620).

^#^Create weight according to month*fsa2*sex*agegroup distribution in school staff sample.

**Supplemental Table 3: SARS-CoV-2 seroprevalence among school staff sample by school district**

|  | **School district** | | |
| --- | --- | --- | --- |
| **Variable** | **Vancouver (n = 1277)** | **Richmond (n = 1277)** | **Delta (n = 1277)** |
| Unadjusted seroprevalence, % [95%CrI] | 23.2% [20.9% - 25.6%] | 24.2% [20.3% - 28.9%] | 25.3% [21.3% - 31.6%] |
| Adjusted seroprevalence, % [95%CrI] | 25.9% [23.0% - 29.0%] | 27.1% [22.5% - 32.6%] | 28.3% [23.6% - 35.6%] |

^&^Teacher, librarian, Student Support Workers; ^#^Principal, Vice-principal, Administrative Assistant; ^@^maintenance staff, school district office staff, or other; *****e.g., work at multiple schools / district office; ^Cumulative incidence of COVID-19 from January 15,2020 to January 27, 2022; ^&^School staff on which VSB had sufficient HR data available; NA = Some data were not be provided by school authorities for the two smaller districts; 10 participants moved to other school districts and therefore have been excluded from this table; 95%CrI: 95% credible intervals.

**May 12, 2022, 8:00 AM**

**APPENDIX 1: COVID-19 mitigations measures in Vancouver schools (2021-2022 school year)**

In August, prior to school opening 2021, the District implemented the 2021-2022 Communicable Disease Prevention Plan including additional COVID-19 Specific Prevention Measures - version 1, September 2, 2021. This document was revised as provincial guidance changed through to March 2022 - version 8: <https://www.vsb.bc.ca/COVID-19/updates/Pages/default.aspx> .

This CD Prevention Plan was based on guidance from the Provincial COVID-19 Health & Safety Guidelines for K-12 Settings:

- The BC Ministry of Education Guidance document: *K-12 COVID-19 Health and Safety Guidelines*: <https://www2.gov.bc.ca/assets/gov/education/administration/kindergarten-to-grade-12/safe-caring-orderly/k-12-covid-19-health-safety-guidlines.pdf>,
- The BC Centre of Disease Control (BCCDC) document: *COVID-19 Public Health Guidance for K-12 School Settings*: <http://www.bccdc.ca/Health-Info-Site/Documents/COVID_public_guidance/Guidance-k-12-schools.pdf>,
- The advice of the regional public health authority, Vancouver Coastal Health (VCH).

The purpose of this document is to lay out communicable disease prevention and control measures, including COVID-19, and address school specific matters as they relate to prevention. The document serves the district’s employees, students, parents/guardians, volunteers, contractors, and visitors by providing appropriate information that can be used to prevent and reduce the risk of contracting and transmitting communicable disease in the district schools and workplaces.

The communicable disease prevention measures and controls included: public health measures (e.g., protocols for testing PCR and Rapid Antigen Tests, contact tracing), environmental measures (e.g., spread out to reduce crowding in classrooms / other school spaces, cleaning and disinfection 1x/day, improved fresh air intake), administrative measures (e.g., reducing crowding indoors - staggered recess/snack, lunch and class transition times, occupancy limits, scheduling appointments for parents/guardians/essential visitors, sign in and out procedures), personal measures (e.g., daily health checks, stay home if sick, physical distancing, hand hygiene, respiratory etiquette), and the use of personal protective equipment (PPE) (e.g. non-medical face masks) by all staff while indoors and for students Gr. 4-12 from school opening on September 7, 2021. This was followed by expanding mask requirements for students K-3 September 27, 2021, by the Vancouver School Board.

Formal Daily Health Assessments were required by all staff and students (via parents) prior to arriving at school and confirmed upon arrival. Anyone with even minor symptoms of cold or flu-like illness was to stay home or go home if these symptoms developed mid-day. Classrooms and other spaces were arranged to maximize distance between students and staff. Class sizes were set by grades: 20 students / class for kindergarten; 22 for grades 1 to 3; 30 for grades 4 to 12. School staff and their students were assigned specific classrooms which were between 75 m^2^ – 83 m^2^ for elementary students (K to grade 7) and 75 m^2^ – 80 m^2^ for secondary students (grades 8 to 12) with larger spaces available for elective courses (e.g., physical education, food studies, metal, woodworking, automotive).

The plan included school schedules for both Elementary students (K-7) and Secondary students (grade 8 to 12) to receive full day in-class instruction. Remote learning was no longer required or an option.

Ventilation measures included refurbishing ventilation and heating systems (HVAC) to ensure proper design operation; scheduling ventilation systems to run 2 hours prior to and after occupancy; increasing outside air component in all systems through louvre adjustments, adding higher efficiency filters (MERV13); and ensuring occupant control over windows and louvres wherever possible to add fresh air flow in spaces.

Other measures included hand sanitizer in classrooms and common areas, directional traffic flow within the schools was controlled and transitioned to regular patterns, provision of plexiglass as provided for certain staff roles where mask wearing was not always an option and reception areas that were public facing, and the training of all staff on the safety plan and protocols. Regular daily cleaning by custodial staff continued and high touch surfaces were cleaned 1x daily or as required. Shared items in classrooms managed by teachers cleaned frequently as well at secondary school, the students were permitted to disinfect equipment. Non-medical face mask use was required Gr. K-12 for all staff and students while indoors at school sites and on school buses. This guidance did not apply if staff or students did not tolerate a mask for health or behavioural reasons. Most enrolled staff did not wear face shields. Face shields were made available to student support staff who work near students with diverse needs and to first aid attendants. As of March 24, 2022, the mask requirement was lifted and students, staff, and visitors could choose to wear, or not wear, masks, face shields or other personal protective equipment in schools and on school buses. Schools and worksites became “mask friendly” and wearing a mask became a personal choice. As of April 4, 2022, plexiglass barriers no longer recommended in alignment with public health recommendations.

SARS-CoV-2 nucleic acid amplification testing (PCR) was available for anyone with symptoms through the provincial health system and advised for students or staff with fever or new symptoms which persisted for over 24 hours. Tests were generally processed within 24 hours, and positive tests were automatically available to public health which investigated can contact traced cases, beginning within 24 hours. Symptomatic close contacts were asked to seek testing; testing was not used to release COVID-19 cases or contacts from isolation on an earlier timeline. Prior to COVID-19 vaccinations, close contacts, including close contacts at school, were isolated for at least 14 full days. Public Health offered COVID-19 vaccination clinics (secondary school sites only) October - November 2021 for those unable to see their health provider or attend their local community clinics. Mature minor consent was required for students in secondary school (under 19) and families in the community were welcomed. Fully vaccinated individuals under 18 years old with mild symptoms and testing is not recommended are to stay home and self-isolate for 5 days and return to school when symptoms subside. For those not fully vaccinated, isolation for at least 10 days until symptoms pass. See: <http://www.bccdc.ca/health-info/diseases-conditions/covid-19/if-you-have-covid-19#self-isolation>

In January 2022, due to Omicron and its subvariants bringing high transmission rates, public health revised their protocols and PCR testing, notification of individuals, and contact tracing ceased. Under the guidance of public health in consultation with the school district, rolling absentee thresholds are used for monitoring the school and grade level absenteeism. High rates of school and or grade level absenteeism over three consecutive days were to be reported to public health for further advisement. Further pandemic supports on January 20, 2022, included Rapid Antigen Test Kits distributed in a phased approach to all employees and students in the K-12 sector. Immunization rates were high, 89% fully vaccinated in the health region (September 2021 to June, 2022). School closures to control transmission were not required during the study period and no schools were required to functionally close due to staff shortages.

Last day of classes was June 29th, 2022, and June 30, 2022 was the last day for staff (administrative day).
